# UKCTOCS Update: Applying insights of delayed effects in cancer screening trials to the long-term follow-up mortality analysis

**DOI:** 10.1101/2020.11.13.20231027

**Authors:** Matthew Burnell, Aleksandra Gentry-Maharaj, Steven J Skates, Andy Ryan, Chloe Karpinskyj, Jatinderpal Kalsi, Sophia Apostolidou, Naveena Singh, Anne Dawnay, Robert Woolas, Lesley Fallowfield, Stuart Campbell, Alistair McGuire, Ian J Jacobs, Mahesh Parmar, Usha Menon

**Author notes:** **Corresponding Author:** Professor Usha Menon, MRC Clinical Trials Unit at UCL, Institute of Clinical Trials and Methodology, University College London, 90 High Holborn, 2nd Floor, London WC1V 6LJ.

## Abstract

**Background:** During trials that span decades, new evidence including progress in statistical methodology, may require revision of original assumptions. An example is the continued use of a constant-effect approach to analyse the mortality reduction which is often delayed in cancer-screening trials. The latter led us to re-examine our approach for the upcoming primary mortality analysis(2020) of long-term follow-up of the United Kingdom Collaborative Trial of Ovarian Cancer Screening (LTFU UKCTOCS), having initially(2014) used the proportional hazards(PH) Cox-model.

**Methods:** We wrote to 12 experts in statistics/epidemiology/screening-trials, setting out current evidence, importance of pre-specification, previous mortality analysis (2014) and three possible choices for the follow-up analysis (2020) of the mortality outcome - (A)all data(2001-2020) using the Cox-model(2014) (B)new data(2015-2020) only (C)all data(2001-2020) using a test that allows for delayed effects.

**Results:** Of 11 respondents, eight supported changing the 2014-approach to allow for a potential delayed effect (optionC), suggesting various tests while three favoured retaining the Cox-model (optionA). Consequently, we opted for the Versatile test introduced in 2016 which maintains good power for early, constant or delayed effects. We retained the Royston-Parmar model to estimate absolute differences in disease-specific mortality at 5,10,15 and 18 years.

**Conclusions:** The decision to alter the follow-up analysis for the primary outcome on the basis of new evidence and using new statistical methodology for long-term follow-up is novel and has implications beyond UKCTOCS. There is an urgent need for consensus building on how best to design, test, estimate and report mortality outcomes from long-term randomised cancer screening trials.

Trial registration: (ISRCTN22488978, Registration date: 6/4/2000)

## BACKGROUND

Randomised controlled trials (RCT) are the cornerstone of the evidence base for clinical management of millions of patients across the world. RCTs evaluating the mortality impact of cancer screening typically involve large numbers of participants followed up over many years, sometimes decades. The general rule in clinical trials is strict adherence to the statistical analysis plan specified prior to unblinding and analysis of outcome data. Sometimes, during continued long-term follow-up of these trials, new understanding based on evidence from other trials and new analytical methods, may require re-evaluation of the analysis plan.

One important example is the accumulating evidence in cancer-screening trials of a delay of several years before a mortality reduction is observed between the screen and control arms[1-3]. Almost all the cancer-screening trials, breast[4-14], prostate, colorectal and lung[15-31] in their graphic representation of disease-specific mortality over time have reported a delayed difference (if present) between screen and control arms(Table 1). Most have an initial time window in the first several years after start of screening during which there is little or no mortality reduction, followed by one in which the reduction becomes evident[2]. However, almost none of these cancer– screening trials have used analytical methods which formally allow for a non-constant effect (non-proportional hazards). All have described the screening effect using relatively simple methods, usually a single Poisson-based rate ratio (RR)[4, 12, 24, 30, 32, 33] or Cox model with a single hazard ratio (HR) estimate[18, 22]. A single HR is only appropriate if the reduction in hazard rates is relatively immediate and constant over time. In screening trials, such estimates cannot reliably describe the changing effects of screening on mortality over time.

**Table 1:**
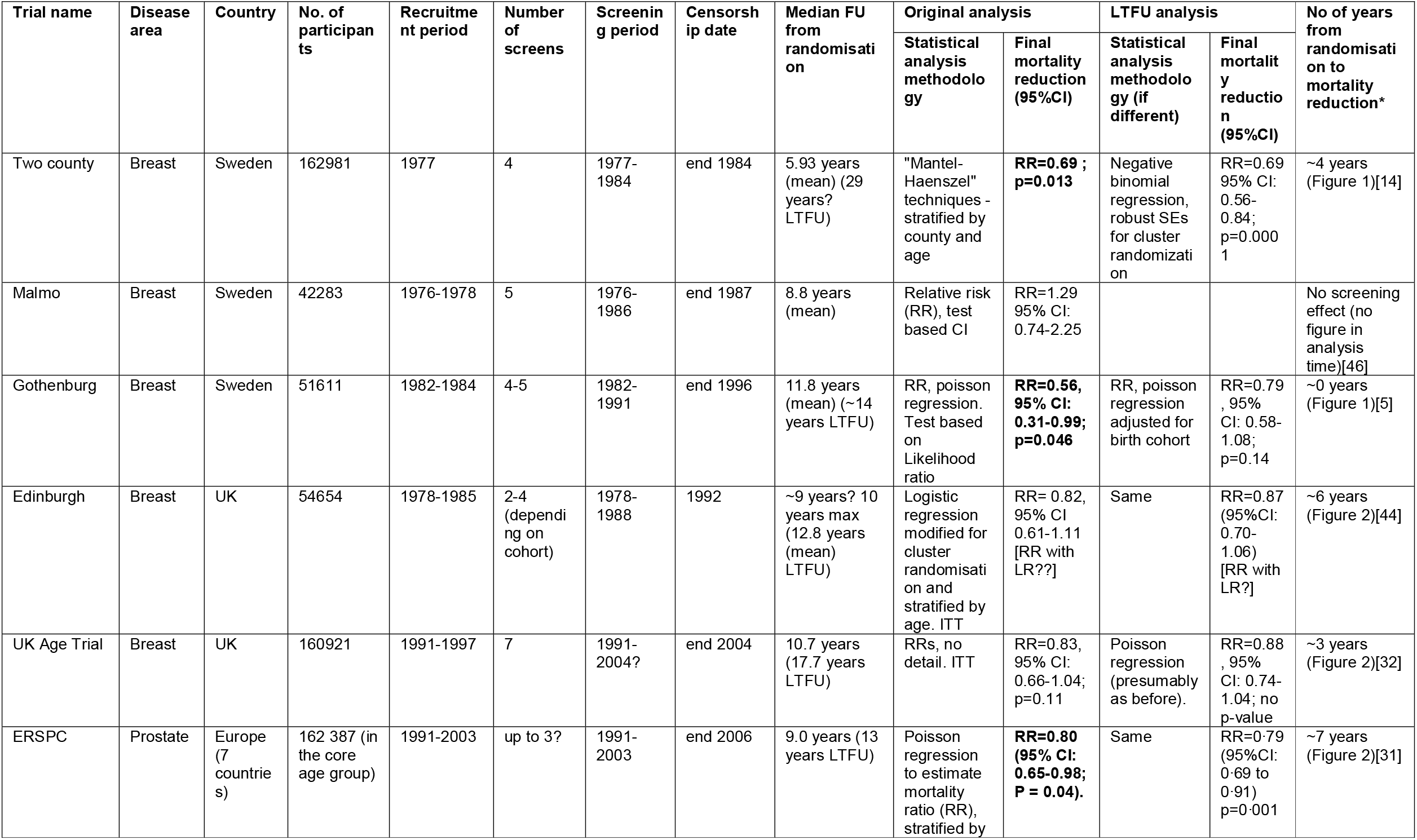
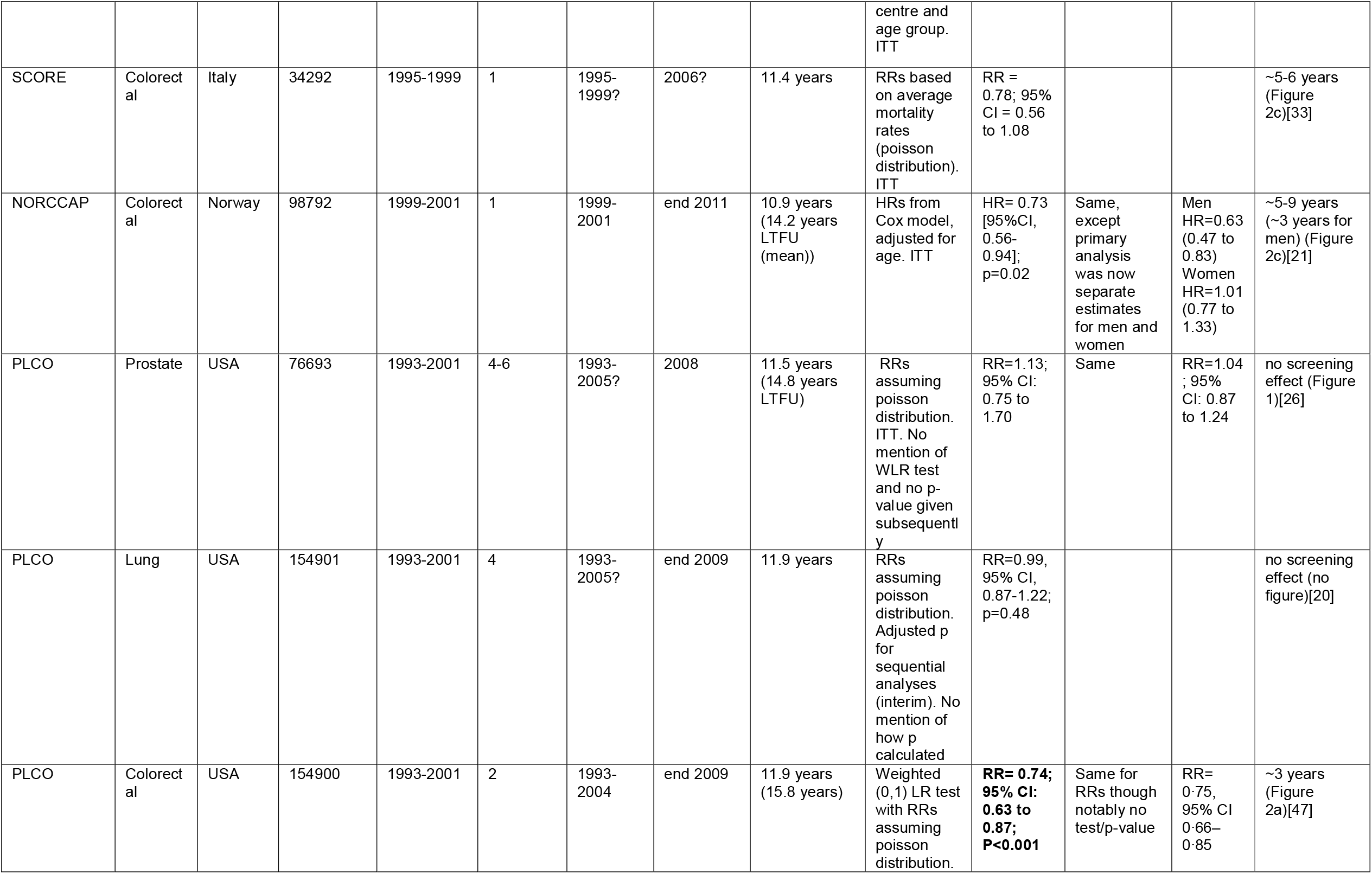
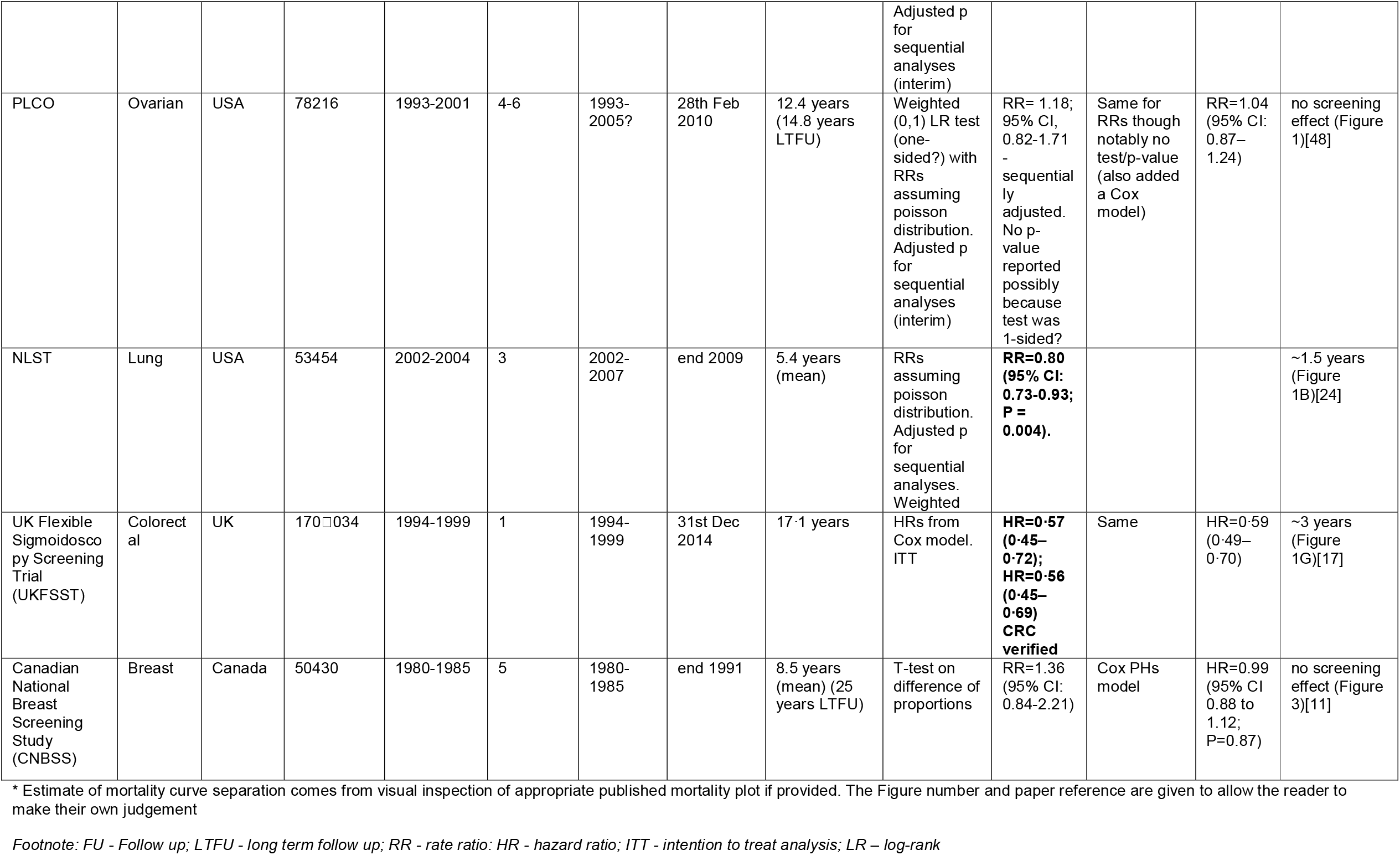
Summary of mortality analyses of randomised controlled cancer-screening trials.

Alongside, new analytical methods have been developed for trials lacking treatment proportionality. Tests that combine evidence from more than one aspect of the data have gained popularity as a way to mitigate the effects of potential but unknown non-proportionality of hazards, although some may work best in a specific scenario. The ‘joint test’ appears in simulations to be preferentially beneficial under late effects[34, 35] whilst the ‘combined test’ appears to be preferentially beneficial under early effects[36, 37]. Another recent addition is the Versatile test[38], which seeks to cover all bases by combining three (weighted) log-rank tests giving good power for the test under early effects, proportional hazards(PH) and late effects, respectively. These tests are likely better suited than the Cox model for analysis of outcomes which are non-proportional across the duration of a trial.

In the United Kingdom Collaborative Trial of Ovarian Cancer Screening (UKCTOCS) too, the initial mortality analysis in 2014 used a PH Cox model and reported an average mortality reduction estimate. However, given the growing external evidence, there have been extensive discussions within the UKCTOCS trial committees to ensure the outcome data is analysed appropriately. We believe that this issue will be important for any long-term cancer screening trial. The Cox model, while valid, could be viewed as restrictive and failing to utilise the most appropriate analytical approach, given the delayed mortality reductions seen in many screening trials across a range of cancers (Table1)[14, 17, 24, 31]. Furthermore, retention of the Cox model based on pre-specification may result in suboptimal interpretation of UKCTOCS data and therefore an abrogation of our responsibility to the huge collective investment by the trial volunteers, the funding agencies, charities, the National Health Service (NHS), researchers and most importantly women who develop ovarian cancer in the future. This is balanced by a concern that changes to the 2014 analysis plan could be controversial and lead to criticism of cherry-picking methodology that gives the ‘best’ test result.

Many trialists may face similar dilemmas, when new evidence suggests that trial design, conduct or analysis may need to be amended. Decisions are often made by the Trial Management Committee (TMC) with input from independent oversight bodies such as a Trial Steering (TSC) or Scientific Advisory (SAC) Committees. We report on the process we undertook in UKCTOCS to re-examine our approach for the upcoming analysis (2020) of the primary mortality outcome at the end of extended follow-up and how we addressed the issue of delayed effects.

## METHODS

Between 2001 and 2005, 202,638 postmenopausal women aged 50-74 were recruited to UKCTOCS. They were randomised to screening using a longitudinal serum CA125 algorithm (multimodal group, MMS, 50,640), transvaginal ultrasound (ultrasound group,USS,50,639) or no screening (control group,C,101,279) as described previously[39-41]. Women in the screen groups underwent screening until the end of 2011 and received a median of nine annual screens. At median follow-up of 11.1 years (administrative censorship 31 Dec 2014), a higher proportion of women were diagnosed with low-volume (stage I, II, and IIIa) tubo-ovarian cancer in the MMS(40%;p<0.0001) compared to C(26%) group. The Cox-model indicated a trend to mortality reduction in favour of MMS (HR 0.85;95%CI:0.70-1.03,p=0.10) and USS (HR 0.89;95% CI:0.73-1.07,p=0.21), which was not statistically significant at the 5% level. A Royston-Parmar (RP) flexible parametric model showed that HR varied over time. In the MMS group, it was 0.92(95% CI:0.69-1.20) in years 0-7 and 0.77(95% CI:0.54-0.99) in years 7-14. In the USS group, it was 0.98(95% CI:0.74-1.27) in years 0-7 and 0.79(95%:CI 0.58-1.02) in years 7-14[39]. Follow-up was extended to 30 June 2020 to assess the long-term mortality impact (LTFU UKCTOCS)[39, 42]. Final receipt of death data from the registries is anticipated by the end of September 2020, with unblinding and analysis planned for November 2020.

To ensure independent input into our statistical conundrum, the TMC proposed seeking the views of a broad panel of international experts with statistical and screening trial expertise who had not been involved in any aspect of UKCTOCS. The process was developed through detailed discussions with the independent members of the TSC. In September 2019, 12 experts (Table 2) were approached by the Trial Statistician for advice. They were sent a letter briefly describing UKCTOCS together with a summary of the current evidence from other cancer-screening trials, importance of pre-specification and our 2014 mortality analysis results. Three potential options for the primary analysis of the extended follow-up data developed with the TSC were described sequentially, each including possible pros and cons, in a neutral manner. These were:

**Table 2:** Summary of choices and additional suggestions if not in concordance with A, B or C of the experts.

| **Expert** | **Expertise** | **Choice** | **Additional suggestions** |
| --- | --- | --- | --- |
| EX1 | Biostatistics, public health | A | Suggests only include cancers diagnosed from period of intervention. |
| EX2 | Biostatistics, clinical trials and cancer research | A |  |
| EX3 | Statistics | A | Ticked 'alternative' but suggested hybrid of A for testing and C for estimation – interpreted as A |
| EX4 | Cancer epidemiology, prevention and screening | Change analysis | Suggested 'number needed to screen'. |
| EX5 | Biostatistics, cancer epidemiology | Change analysis | Did not complete form but indicated choice by email, test based on difference of restricted mean survival time (RMST). |
| EX6 | Biostatistics and epidemiology | Change analysis | Suggested splitting data into yearly bins and assess HR in each, possibly with smoothing. Avoid single HR. |
| EX7 | Biostatistics, clinical trials and cancer research | C | Did not complete form but indicated choice by email. Prefers more parsimonious model with interpretable parameters. |
| EX8 | Biostatistics, clinical trials | C |  |
| EX9 | Biostatistics, public health | C | Prefers more parsimonious model with interpretable parameters. |
| EX10 | Cancer epidemiology, public health | C | Also suggests 'versatile weighted log-rank test' |
| EX11 | Statistics, public policy | C |  |
| EX12 | Biostatistics | - | Did not respond within timeframe |

A. analyse all outcome data (2001-2020) using the PH Cox-model of the original UKCTOCS analysis, representing the pre-specification viewpoint
B. analyse only the outcomes that occurred since the original censorship (31 December 2014), either assuming PH or not, to address the view that data should not be re-used, without formal statistical accommodation for multiple analyses.
C. model all outcome data using a method of analysis and model that allows for a late effect of screening on mortality and reflects current understanding of cancer-screening trials - a pragmatic evidential approach. The specific model suggested for C) was the RP model[43] as it had been used as a secondary analysis method for the 2014 analysis[39].

Experts were asked to critique and state a preference or suggest another option (Supplementary Materials 1). Results were collated and summarised based on 1) indicated choice of A, B, C or other and 2) pertinent comments provided.

## RESULTS

In total 12 individuals were contacted from the UK (5), USA (5), Canada (1) and Belgium (1) and 11 responded (see acknowledgement). Their anonymised responses can be found in Table 2 and Supplementary Table 1.

Eight (73%) of the 11 experts recommended changing the pre-specified analysis to one that more appropriately allows for a delayed effect (Table 2). *EX4* was not troubled by the shift from a pre-hoc to post-hoc decision - “reason” should have a role in science. Similarly, *EX8* argued “a conclusion should be reached based on a proper consideration of the full evidence” and use scientific principles – “full information from data should be extracted”. Indeed, rather than viewing it as “data-dredging” or “changing the endpoint”, *EX8* described this approach as just “using common sense”. *EX9* felt the lack of (complete) pre-specification a weakness, but not “a violation of good scientific principles”. For “a major and definitive screening trial ….. such regulatory constraints should not be the primary consideration” but instead “approximating the truth as well as possible”. *EX11* was not persuaded by the pre-specification argument, and claimed keeping a plan that is less preferable “turns research rules into an irrational, mindless, and restricting obsession with methodological procedure”; “rules have a purpose, but when the higher priority is understanding phenomena in a reasoned disciplined way… then a compelling argument can be made to deviate from them”. *EX11* stated that no screening trial has shown an immediate effect and appealed to the common sense of the scientific audience; “we can discern the difference in attempts by a study team to game the analysis to gain statistical significance, from a good faith effort to apply a statistical technique that is more appropriate for the data”. Different screening trials will have different results and delayed effects, all dependent on differing facets of trial design and the cancer itself, the effects of which are largely unknown until we do the study. “Point is, we are still learning how to design and analyse RCT screening trial data.”

Three of the eleven (*EX2, EX3, EX1*) believed that we should retain the initial analysis approach (option A). This was based on the pre-specification argument - “avoids the appearance of trying to get a significant result by changing the test”(*EX2)*, “maintains credibility in the scientific community”(*EX3*), “most likely to be accepted as valid by the cancer research and policy community”(*EX1*). However, *EX1* did suggest modifying the pre-specified plan to limit analysis to only cancers diagnosed within the screening period.

Of the eight who suggested changing the pre-specified analysis, five (*EX7, EX8, EX9, EX10* and *EX11*) explicitly selected approach C (using all acquired outcome data and a model that allows for delayed effects). While there were positive comments about the suggested RP model (credibility due to pre-specification *EX7*, informative of the screening effect over time *EX9)*, none gave a clear endorsement of this approach. The main reason was interpretability (*EX7, EX9, EX4, EX6). EX10* noted that power was little studied under various “flavours” of non-PHs, and suggested separating testing from estimation, opting for a versatile weighted log-rank test for the former. *EX4* and *EX6* formally indicated an alternative option. *EX6*’s preference was for dividing the data into yearly bins and estimating the HR in each, possibly with some smoothing. *EX6* argued extensively we should avoid a single HR estimate, which will provide “a very blurred, incomplete and misleading picture of how much/little good screening did for the 100,000 participants screened, or of how much future women might expect from a screening regimen based on these screening tools.” *EX4* stated that the number needed to screen was the most suitable measure for a screening study. *EX5* recommended a test based on the difference of restricted mean survival times (RMST) which “does not need any modelling and the results can be interpreted easily clinically”.

None of the 11 responders chose Approach B. This was mainly because it did not use the full dataset. In addition, there were concerns that it could lead to ‘unfavourable early results (important data) being censored(*EX11)* and a “disconnected” HR*(EX6)*.

Based on the feedback, we decided to change the primary analysis test for LTFU UKCTOCS. Table 3 summarises the major pros and cons of available approaches to dealing with non-PH in terms of tests. We used two main criteria to choose the specific test - (1) minimal *a priori* specification on the specific form of the mortality difference over time (2) able to accommodate delayed effects while maintaining good power in a variety of potential scenarios. Based on these criteria, we opted for the Versatile test[16], suggested by *EX10*. The RP model was retained to estimate absolute differences in disease-specific mortality at 5, 10, 15 and 18 (our estimate of the upper limit of reliable follow-up given administrative censorship on 30 June 2020) years. Options A and B were included as secondary analyses of the primary mortality outcome. These amendments were incorporated into the statistical analysis plan (20 February 2020), which was endorsed by the independent TSC.

**Table 3:**
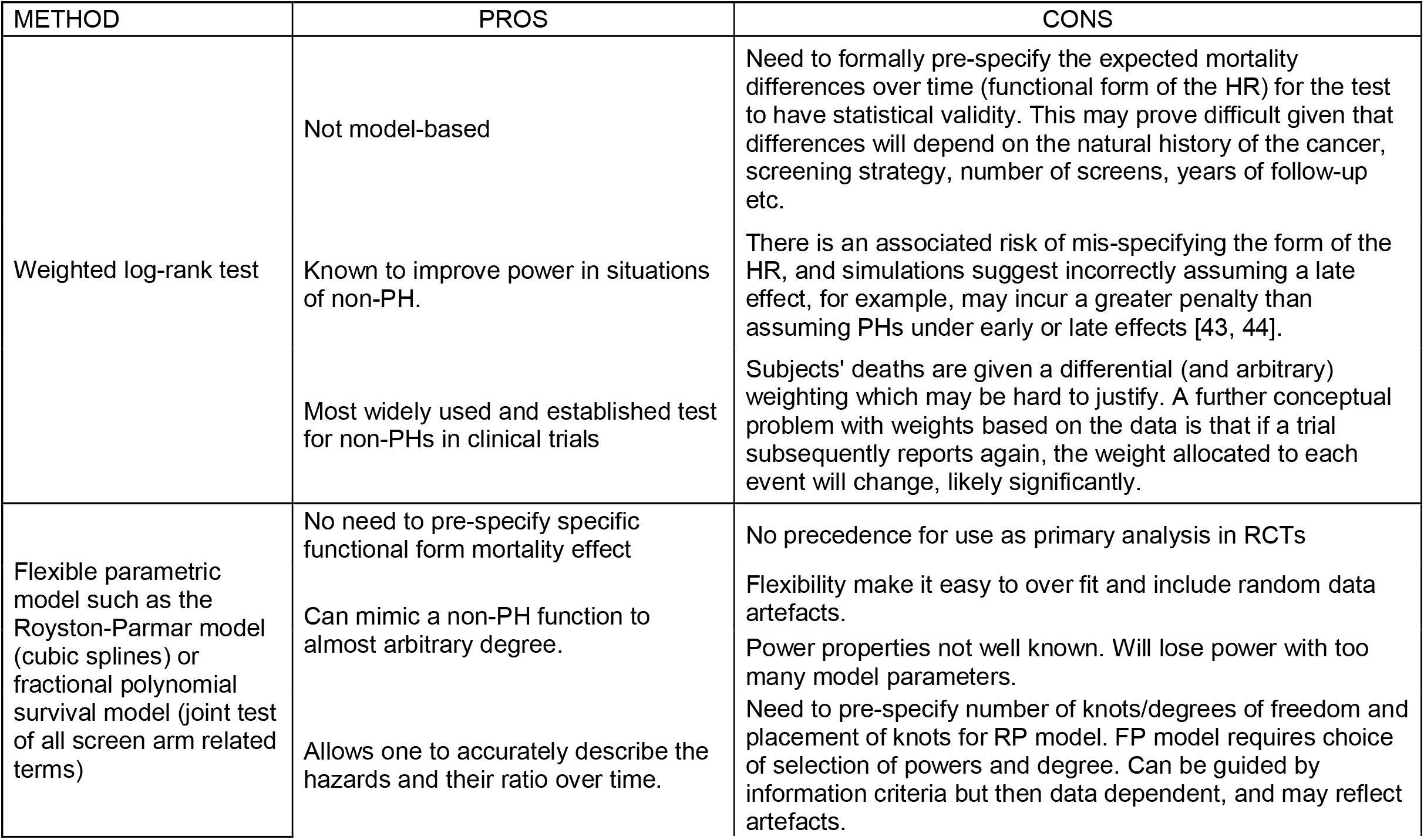
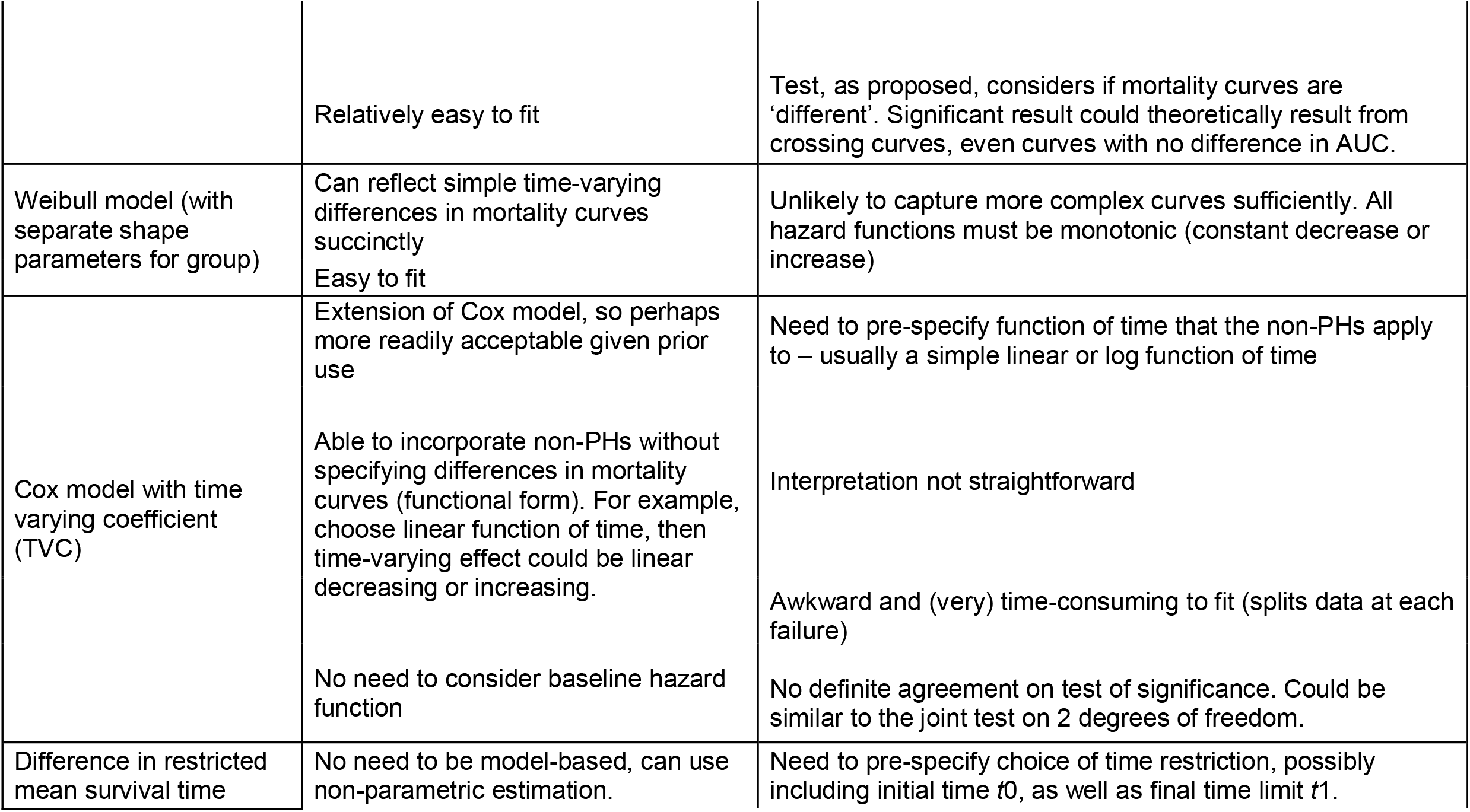
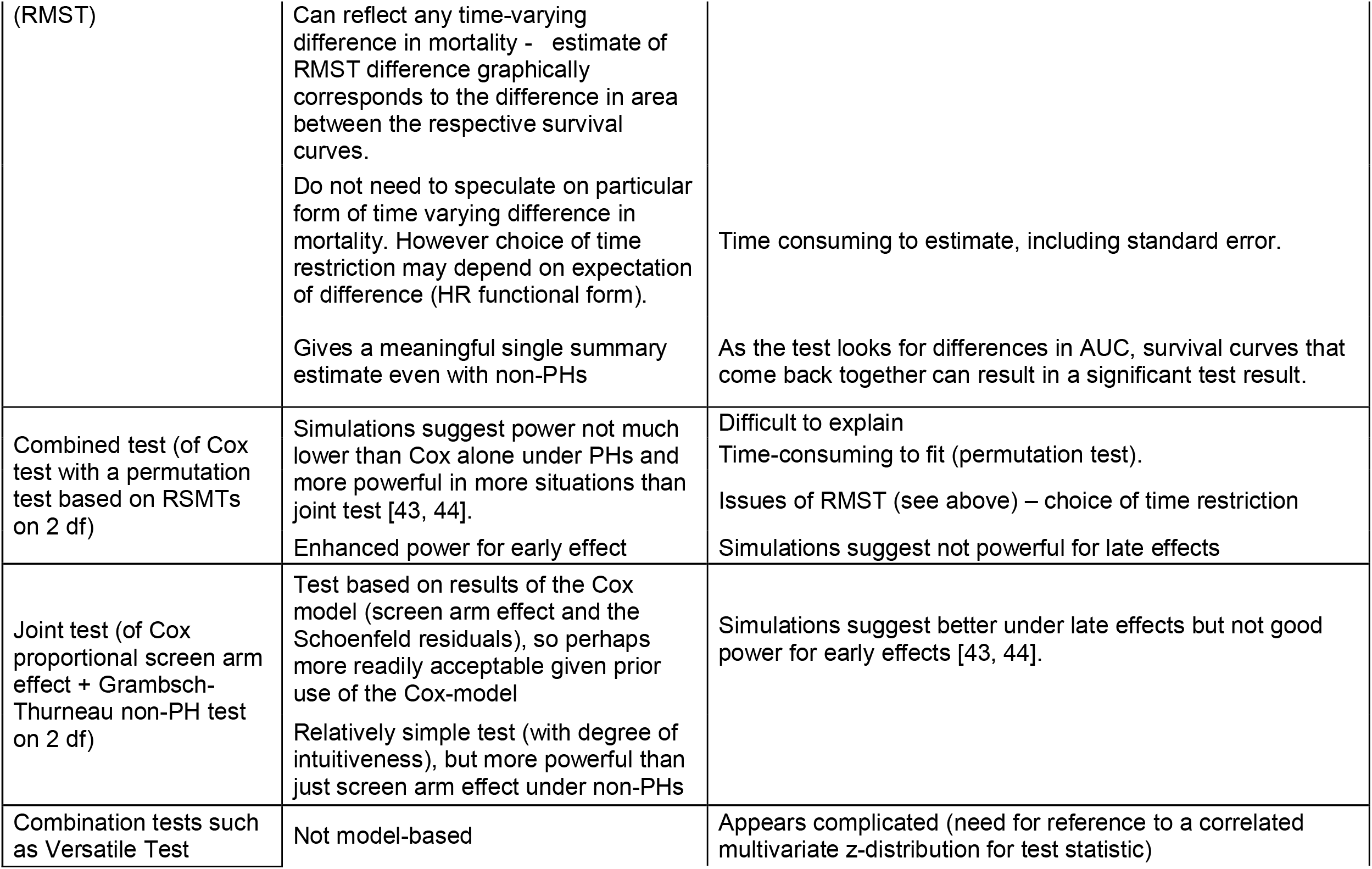
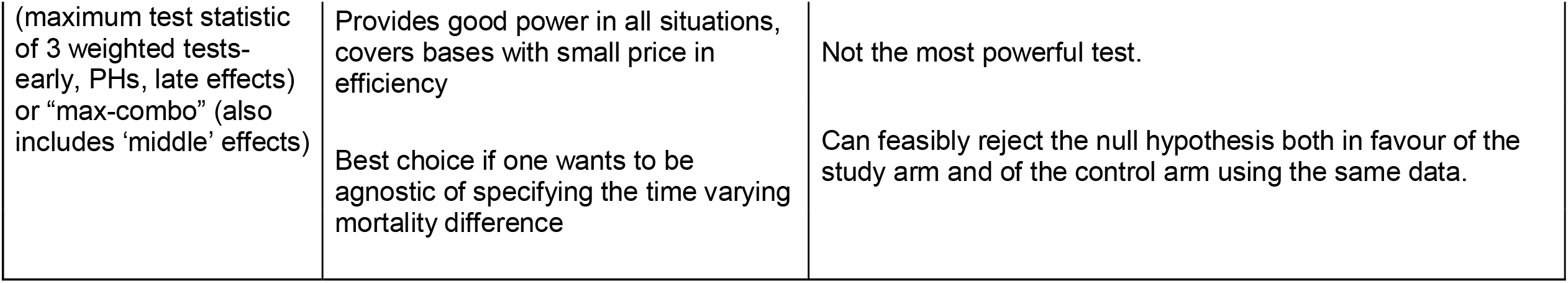
Summary of pros and cons of potential statistical tests that could be used when there is a time varying mortality difference (non-proportional hazards)

## DISCUSSION

Given the now large body of evidence of a delay in mortality reduction in long-term cancer-screening randomised trials, and the majority view of independent statistical, epidemiological and screening trial experts, we altered the approach for our primary mortality analysis for the LTFU from that used for our 2014 analysis. The new approach allows for a delayed effect in contrast to our previous analysis which assumed a constant screening effect. There were a variety of opinions on the specific test which suggests an urgent need for consensus building on how best to design, analyse and report mortality outcomes in cancer-screening trials.

Our decision to change the statistical analysis plan for extended follow-up is a significant decision. The large majority of the published cancer-screening trials[17, 25, 26, 31, 32, 44] have retained the same primary mortality analysis methodology for both their initial and extended follow-up analysis (Table 1). The only exceptions we found were the Two County trial which used negative binomial regression[14] for follow-up analysis in place of Mantel-Haenszel stratified risk-ratios[12] and the Norwegian Colorectal Cancer Prevention Trial (NORCCAP) which changed the primary analysis from overall population to subgroups based on gender[21]. In the Two Country trial, whilst no explanation was given, the change was not substantive; both initial and follow-up methods estimated risk ratios. For NORCCAP, “because substantial heterogeneity existed between women and men, the steering committee decided to present results for women and men separately”, which may be argued as a significant post-hoc data-driven amendment. None of the trials as far as we are aware sought independent expert opinion. In contrast, we undertook an external consultation. Although the independent expert panel was not unanimous, the majority concluded that a rational argument for revision outweighs that of procedure and pre-specification, and recommended choosing the most appropriate test that allows for a delayed effect. We accepted the view of *EX7* that one should “do what you yourselves think is the most effective and secure analysis of all your data, bearing in mind the current state of information about the field.” There will be debate about our decision, which we welcome, given the broader implications.

A number of factors contribute to delayed mortality effect. In the early trial-years, the absolute death rates are low as a result of eligibility criteria which exclude women with cancer diagnosis. The time interval for an individual to be diagnosed with cancer after joining the trial and then dying of the disease also contributes to the delay in separation of the mortality curves. Additionally, the impact of screening on cancers detected at the initial prevalence screen is reduced, as these are necessarily more advanced when screen-detected compared to screen-detected cancers in later years. The performance of most screening strategies improve over time as the number of screens accumulate and the teams involved get more experienced. This is magnified when longitudinal biomarker algorithms are used as they are based on detecting change from baseline. Finally, the length of follow-up after end of screening impacts on the specific form of the mortality difference over time as the longer the interval, the greater the dilution of screen-detected cancers by cancers that develop after the end of screening[32].

The PLCO colorectal[29] and ovarian[19] trials used a test that has better power for the delayed effect described above. Both used the weighted log-rank test, which is perhaps the best known method for improving power in such situations. However, it requires correctly anticipating the specific form of the mortality difference over time, which will depend on the natural history of the cancer, screening strategy, number and frequency of screens and years of follow-up. We have chosen the Versatile test[38], introduced in 2016, which does not require pre-specification of the mortality difference over time. It combines three (weighted) log-rank tests appropriate for capturing early effects, PH and delayed effects, respectively. It is therefore versatile enough to maintain good power in all potential scenarios, rather than optimal in any given scenario.

Unlike other trials, including the PLCO colorectal[29] and ovarian[19] trials, who measured the screening effect using a single ‘averaged’ rate-ratio, we will use a flexible parametric model to estimate absolute differences in disease-specific mortality at 5,10,15 and 18 years. This is in keeping with the growing view that to adequately describe what might be achieved with a particular cancer screening strategy, a more comprehensive set of time-specific measures needs to be reported. Hanley *et al* has extensively re-analysed cancer screening trial data and shown that a one-number summary measure systematically dilutes the estimate of mortality reduction that results from screening[2]. In the most recent re-analysis involving breast cancer screening data from Funen, Denmark, the average mortality reduction was 18% using a PH model and ranged from 0 to 30% when a non-PH model was used that considered the impact at different points over time. The reductions were largest for periods where sufficient time had elapsed for the impact to manifest[45].

The key strength of our approach is the independent and transparent process we have adopted to address a challenging issue and the criteria we used to choose a new specific approach. This involved accommodating delayed effects while maintaining good power in a variety of potential scenarios and requiring minimal *a priori* speculation on the specific form of the mortality difference over time. A limitation is that given the orthodoxy surrounding pre-specification for analysis of trials, we have retained the original Cox model with an averaged HR over time as an estimate for our secondary analysis.

The screening community is only beginning to understand the challenges posed by long-term cancer-screening trials. Mortality reductions may have been underestimated across cancer types by not considering their timing. Given the importance of early detection in many national cancer strategies, we hope our report will accelerate much needed consensus building on how best to design, analyse and report trials testing cancer screening strategies – as it is clear our currently accepted and widely used methods are insufficient. We also hope it will encourage debate and transparency on how advances in understanding and new analytical methods can be evaluated and incorporated into long-term trials.

## Data Availability

Tables 2 and Supplementary Table 1 contain comments provided by the experts.

## List of abbreviations

(UKCTOCS): United Kingdom Collaborative Trial of Ovarian Cancer Screening
(LTFU UKCTOCS): Long-term follow-up of the United Kingdom Collaborative Trial of Ovarian Cancer Screening
(RCT): Randomised controlled trial
(RR): Rate ratio
(HR): Hazard ratio
(CI): Confidence interval
(PH): Proportional hazards
(TMC): Trial Management Committee
(TSC): Trial Steering Committee
(SAC): Scientific Advisory Committee
(MMS): Multimodal group
(USS): Ultrasound group
(RP): Royston-Parmar model
(NORCCAP): Norwegian Colorectal Cancer Prevention Trial
(PLCO): Prostate, Lung, Colorectal and Ovarian Cancer Screening Trial

## Declarations

### Ethics approval and consent to participate

The initial study was approved by the UK North West Multicentre Research Ethics Committees (North West MREC 00/8/34) on 21 June 2000 with site-specific approval from the local regional ethics committees and the Caldicott guardians (data controllers) of the primary care trusts. The long-term follow-up amendment was approved on 24 January 2017 and the amended protocol including the new statistical plan was approved on 12 May 2020. All trial participants provided written informed consent.

## Consent for publication

All authors have seen the final version of the manuscript and give their consent for publication.

## Availability of data and materials

Tables 2 and Supplementary Table 1 contain the exact comments provided by the experts.

## Disclaimer

The views expressed are those of the authors and not necessarily those of the NHS, the NIHR or the Department of Health and Social Care.

## Author contributions

The process was conceived following many discussions within the TMC involving all authors. MP and UM supervised the study. MB performed the literature search. MB, SJS, AMcG, and MP proposed the statistical analysis options with further input from JC (TSC). The survey was drafted by MB, AGM, MP and UM with input from IJJ, AMcG, and SJS. AGM, AR and MB collated the results and MB undertook analysis. All contributed to data interpretation. MB prepared the tables. MB, AGM and UM drafted the manuscript. AMcG, LF, SA, JK, RW, IJJ, MP and SJS helped revise the draft. All authors critically reviewed the manuscript and approved the report before submission.

## Acknowledgements

We are hugely grateful to the international panel of experts (Professor Marc Buyse, Professor David Cox, Professor Stephen Duffy, Professor Mitch Gail, Professor Jim Hanley, Professor David Harrington, Professor Patrick Royston, Professor David Schoenfeld, Professor Robert Smith, Professor David Speigelhalter, Professor LJ Wei) who have contributed their time and expertise. We are also indebted to the insights and support provided by the members of the Trial Steering Committee - Professor Henry Kitchener (Chair), Professor Julietta Patnick, Professor Jack Cuzick and Ms Annwen Jones. We thank all 202,638 volunteers without whom the trial would not have been possible and all the staff involved in this trial for their hard work and dedication.

## Supplementary material legends

**<u>Supplementary Material 1:</u>** Cover Letter to Independent International Expert panel, Outline of Options, Comment Form

**<u>Supplementary Table 1:</u>** Summary of Responses from Independent International Group

