## Supplementary materials for "UKCTOCS Update: Applying insights of delayed effects in cancer screening trials to the long-term follow-up mortality analysis"

Burnell M et al

#### Supplementary Material 1

#### Cover Letter to Independent International Expert panel

Prof [to insert]

Address [to insert]

23rd September 2019

Dear Prof [to insert],

We are writing to you for your views on an important statistical issue, which has arisen in the large, long-term UK Collaborative Trial of Ovarian Cancer Screening (UKCTOCS) which has now been running for 19 years.

**Brief Overview of the trial**

The UK Collaborative Trial of Ovarian Cancer Screening (UKCTOCS) began recruiting in 2001 and aimed to be a definitive RCT on ovarian cancer screening in having sufficient size and duration to investigate ovarian cancer mortality as the primary endpoint, while comprehensively addressing the other secondary end points such as compliance, morbidity (physical and psychological) and cost effectiveness.

Between 2001-5, 202,638 normal-risk post-menopausal women were randomised through 13 trial centres to annual screening using a multimodal screening (MMS) or ultrasound screening (USS) strategy or no screening in a 1:1:2 ratio. Screening (involving 673,765 annual ultrasound and blood tests) continued until the end of 2011 with women receiving a median of 9 and a maximum of 11 annual screens. Follow-up included electronic health records linkage to cancer and death registry and postal questionnaires.

MMS used serum CA125 measurement, with significant increases above a woman's own baseline detected by the Risk of Ovarian Cancer Algorithm (ROCA). ROCA estimated the risk of having ovarian cancer given the woman's age and CA125 results, and triaged women to normal (annual screening), intermediate (repeat CA125 testing in 3 months), and elevated (repeat CA125 testing and transvaginal USS as a second-line test in 6 weeks) risk.

Annual screening in the USS group used transvaginal USS as the primary test, which was classified as normal (annual screening), unsatisfactory (repeat in 3 months), or abnormal (scan with a senior ultrasonographer within 6 weeks). In both groups, women with persistent abnormalities had clinical assessment and additional investigations within the NHS by a trial clinician. Screen positives were those who were recommended to undergo biopsy/surgery because of a suspicion of ovarian cancer.

Mortality from ovarian cancer (ICD 10 codes C56 and C57.0) by 31st December 2014 was defined as the primary outcome measure.

#### Brief overview of the Statistical Issue

The Statistical Analysis Plan (SAP) specified a Cox test of each screen arm versus the control arm for the primary analysis, with a Dunnett-style correction. The Primary Results (as we considered them at the time) were published in the Lancet in 2016 (Jacobs Menon *et al* Lancet 2016; 387: 945–56)[1].

Although a delay in the mortality effect of screening had been pointed out by others who noted that "it does not obey the proportional hazards assumption" (Warwick & Duffy)[2], no mention was made of anticipating a delayed effect in the Statistical Analysis Plan for UKCTOCS. The delay in UKCTOCS was about half the duration of the study, as the Kaplan-Meier (KM) curves appeared to show no discernible difference in mortality until about 7 years into the study.

**Figure 1:** Kaplan-Meier failure curves of ovarian cancer mortality (per 100 000 women). *From Jacobs Menon et al Lancet 2016; 387: 945–56*[1]

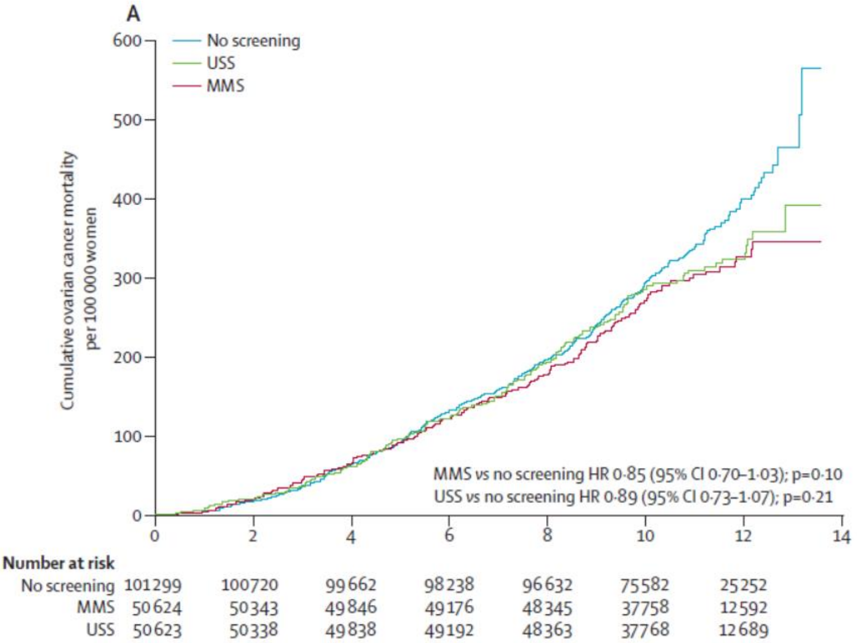

The mortality reduction, as measured by the hazard ratio (HR) in the Cox model, was 15% for MMS (p=0.10) and 11% for USS (p=0.21). HR estimates from a Royston-Parmar model showed this mortality effect was made up of a small effect (8%) in years 0-7, and a larger effect 23% in years 7-14 for MMS, and 2% and 21% respectively for USS. This (apparent) delay in the mortality effect occurs in all screening trials where a mortality effect has been detected, the delay ranging from a few years to ≥7 in two of five such trials reported during UKCTOCS. Thus the default for screening trials should be a delayed effect with immediate effects including proportional hazards forming the alternative hypothesis. [However, it should be noted that were proportional hazards to be expected and thus form the null hypothesis, tests of non-proportional hazards would not be conventionally statistically significant at the 5% level with p=0.095 for MMS and p=0.108 for USS using the Grambsch-Therneau test of Schoenfeld residuals in the unit-timescale]. The results indicated that there might be a significant screening effect if further follow-up were allowed, providing more information beyond the point of the apparent delayed mortality reduction. Therefore, a statistical model which allows for a delayed effect, and thus non-proportional hazards, is likely to be a more powerful test of statistical significance than a test which is most powerful under proportional hazards.

Given the highly significant difference in early stage ovarian cancer between the MMS and control group on an intention to screen analysis, the ambiguous nature of the result, that there are no other large trials of the MMS screening strategy as used in UKCTOCS and the likelihood that such a large and long term screening trial in ovarian cancer will not be repeated, funding and ethical approval was secured in order to extend follow-up for a further 5-6 years, dependent on Control arm events. The hope is that this will provide a definitive conclusion on screening for ovarian cancer.

The end of this follow-up period will be during 2020, and so the need to finalise and sign-off on the Statistical Analysis Plan is now necessary. However, in these circumstances there are pros and cons as to the most appropriate primary analysis to be employed. The choice is inevitably complicated by the fact that we have done and reported one analysis of this trial already. Three possible general approaches have been proposed as follows.

**A.** To continue with a Cox/logrank test on all the accumulated data

**B.** To use a model and test on all the accumulated data that allows for a delayed effect

**C.** To perform a Cox test on just the 'new' data (2015-2020) acquired since censorship at the end of 2014 in the original analysis

Each of these approaches has its strengths and weaknesses. In the rest of this document we describe these 3 methods, their pros and cons as far as we have considered them.

We would welcome your views of which (if any) of these approaches you would consider the most appropriate in our situation and why.

Once we have received responses from all statistical experts, we shall summarise these and present a consolidated view to our executive Trial Steering Committee.

We would be grateful if you could respond by the 11th October please.

Yours sincerely,

Prof Max Parmar *DPhil, OBE* & Prof Usha Menon *MD, FRCOG*

on behalf of the UKCTOCS trial investigators

### Option Outlines

#### APPROACH A: To continue with a Cox model fitted to all accumulated data

##### 1. What is the method?

Approach A analyses all the data generated from 2001 to 2020 in the same manner as the original analysis, using a Cox/logrank test which is most powerful under proportional hazards.

##### 2. Why is this a good method to use?

The principal benefit to Approach A is the continuity of the Statistical Analysis Plan, and the appearance that methods have not been altered on the basis of the observed results to date, simply to provide the most significant result. The Long-Term Follow-Up (LTFU) of UKCTOCS is only running due to the ambiguous nature of the result of the original primary analysis; a clear positive or 'negative' result is unlikely to have led to grant applications and further follow-up. Altering the method of analysis once results have been previously observed may lead to criticism from the scientific community, particularly that the p-value has lost any real interpretability.

Results from other screening trials, where there was an effect, all showed a delay. Additional UKCTOCS data may further confirm this delayed effect and resultant non-PHs, but we do not know for sure that an alternative method would prove more powerful. The Cox model can still be powerful (relative to the most powerful test) under modest deviation from PHs.

Other general benefits include the simplicity of the model and not needing to specify the baseline hazard.

The literature for screening trials has produced multiple articles that report on longer-term primary outcomes, sometimes many years after the first mortality analysis. For example the HIP[3], the Stockholm trial[4], the Edinburgh trial[5], the Swedish 2-country trial[6], the Canadian NBSS[7], the UK Age trial in breast cancer[8]; the Swedish RPCS trial[9], the PLCO in prostate cancer[10]; the UK Flexible Sigmoidoscopy trial[11], the PLCO in colorectal cancer[12]; and the PLCO in ovarian cancer[13] have all reported again with extended follow-up, and all without any mention of adjustment to alpha or issues associated with re-analysing data. Importantly, in all these trials there was no or very little change to the analysis methods - and all have assessed mortality reduction using simple hazard-ratios or even risk-ratios.

One trial of particular pertinence to UKCTOCS is the European Randomized Study of Screening for Prostate Cancer (ERSPC). They first reported a marginal result (HR=0.85; 95% CI:0.70-1.03 after 9 years FU[14]) and then re-used the extant data with freshly accumulated data to report on a fuller picture of the overall screening effect (HR=0.78; 0.66-0.91) after 11 years[15], and 0.79 (0.69-0.91) at 13 years.[16] Their trial also observed around 7 years delay before separation of the mortality curves became apparent. The respective shape of these curves did not substantively change in the two further reports - however many more events were observed in the later period where the mortality had separated, giving greater credence to a late effect rather than no effect. In these 2 later papers the authors used the same methods of analysis at the primary and later analyses but made no adjustment to their significance testing procedure, although they did acknowledge the issue when using the justification "No adjustment of significance for α-spending in sequential analyses was applied because the present analysis is protocol based and not driven by statistical significance".

##### 3. What are some of the potential criticisms/downside of this method?

One major criticism is that a Cox test is unlikely to be the most powerful method we could choose, and quite possibly far from the most powerful, dependent on the nature of the long term follow-up data. The original Kaplan-Meier curves pictorially suggested separation, even if the statistical evidence was not as persuasive. However, if the LTFU data fills in the picture in a similar manner to the extant KM curves then non-proportionality should become quickly evident, and the Cox model would be sub-optimal in terms of both power and reflecting reality. A simple interpretation, merely to follow convention, becomes harder to justify as a benefit. In addition, since the FU period is entirely after (and as much as 8 years after) the end of screening, then there is concern the proposed screening effect will start to become diluted; even allowing for a considerable lag time that, in theory, should mirror the initial observed delayed effect. Dilution would impact the likelihood of a positive result; it would also be another source of non-PHs (this time convergent curves) that a Cox model would ignore.

#### APPROACH B: To model just the 'new' data acquired since censorship of the original analysis (2015 onwards)

##### 1. What is the method?

This approach will ignore the data that has been previously analysed, and start the analysis point from the end of censorship of the original analysis. That would mean follow-up time ranges from 1st January 2015 until the point in 2020 when the scheduled number of events have been reached (232 new Control arm events). Only ovarian cancer deaths in this period will contribute to the primary analysis.

The specific statistical method is still to be decided. It is likely that the Cox test is now the most powerful method (or close to) as the LTFU period is entirely after the KM curves appear to diverge (start of FU at minimum $t$=9.2 years, median $t$=11.1 years), although note the dilution effect mentioned in Approach A may affect proportionality toward the end of analysis time. A "combined test" (of a Cox test with a permutation test based on restricted mean survival times on 2df) which is not notably less powerful than a Cox model alone when PHs are true, but enhances power for early effects, may prove a viable alternative.

There is also the choice of the analysis timescale to consider (see point 3.)

##### 2. Why is this a good method to use?

This may be viewed as a constructive option, as a 'clean break' from the original analysis. The data is kept separate from results that have already been published. To mix the 2 datasets introduces awkward statistical issues, when the second analysis is dependent on the result of the first analysis, but was never originally planned for.

The cutting of data from 2001-2014 appears drastic, indeed damaging to the chances of a positive result. However, this is a matter of perspective and it could be argued that Approach B will give a better chance of an '*accepted*' significant result for ovarian cancer screening. Approach B only uses data (an expected 232 control events) from beyond the first 9 years post-randomisation, the first 7 of which showed no difference between the arms with the mortality curves almost superimposable. Based on a Cox model/log-rank test, the power for a significant screening effect on the new data alone compares well to a conditional (on the original observed data) power calculation that uses the same hypothesised screening effect from 2015 onwards.

A benefit of Approach B is that, as a new analysis, we have arguably more freedom to choose the specific method, with less external pressure perhaps to preserve the model used previously. However, if the PH assumption is approximately valid, then the Cox test could be more powerful than a test that assumes a violation, certainly if the latter employs more degrees of freedom (e.g. combined test or a flexible parametric model).

##### 3. What are some of the potential criticisms/downside of this method?

Approach B might be widely viewed as a 'waste' of data where the majority of events in a large, expensive RCT are discarded. The other viewpoint might counter that this data has resulted in a published trial result, and so were very much made use of.

There may be resistance simply because it is a significant alteration to the trial and there may be suspicion over the motives for that change. Furthermore, we could find no precedent for a screening trial adopting this strategy, even though many screening RCTs continue to report on long-term outcomes. The appearance that the original trial has seemingly been diminished to a hypothesis-generating study could prove difficult to accept for many involved.

One peculiarity of starting analysis at 1st January 2015 though, is that a few ovarian cancer deaths that occurred before 2015 but were not known about by mid-2015 (when data was frozen for the initial analysis) will not feature in either analysis.

Analysing from 2015 onwards will only represent a partial view of the screening effect over time - inference will be solely based on late effects, after screening has finished. However, whilst the primary analysis is based on a statistical test of the 'new' data only, other complementary analyses can still utilise the complete dataset and report on hazard rates and effect sizes over time to give the fuller scientific picture.

A delicate issue mentioned in point 1 is the timescale. The original analysis was founded on time from randomisation. To preserve this timescale would require delayed entry, so that some women enter at 9.2 years, up to some entering at 13.6 years in analysis time. In practical terms this may result in odd-looking KM survival curves, and will mean that information will be thinned out over the 9-11 year-period when only a reduced sample of women will enter the study. An alternative is to have all volunteers starting at the same time-point, which can be set at zero. This fixing would also have a universal interpretation, as 3 years on from end of screening - although note there would be a differential time lag to last actual screening episode. Aligning the women's FU period in this way would also mean that any dilution effect will also occur at a common point in the timescale, which was not originally the case. However, the overall effect of this timescale choice may be to diminish the interpretability of the primary result.

#### APPROACH C: To use a model and test on all accumulated data that allows for a delayed effect

##### 1. What is the method?

Approach C analyses all 19 years of mortality data from 2001 to 2020 with a model that allows for a delayed effect. The Royston-Parmar model[17] is a flexible parametric model estimating the cumulative hazard with cubic splines for each arm of the trial. The test for a mortality effect is the multivariate Wald test for the difference between the coefficients of the two splines - as specified by Royston and Parmar.[17]

##### 2. Why is this a good method to use?

**Perspective:** The perspective of this approach owes much to David Cox's article entitled "Statistical Science: A Grammar for Research".[18] In this framework, statistical thinking is fruitful when merged seamlessly with the subject matter in contrast to routine application of formal statistical rules, including even highly regarded rules such as randomization and pre-specification of statistical analyses. Rules are useful but are to be placed in the service of the science, which may lead to not using a rule in a given study. Cox makes the case that while planning a study should include planning the analysis, the analysis may be changed if the data show the pre-specified model is incorrect, or if there is a long time between the planning and measuring the endpoint and during this time more appropriate analyses may have been developed or further information arisen. This is the case with UKCTOCS where 14 years elapsed between the start of the study and availability of the mortality endpoint for the first mortality analysis. During the conduct of UKCTOCS, reports from the five early detection trials with a mortality effect all show a delay. To seamlessly merge with the science of early detection the statistical model should allow for this delay, ruling out the proportional hazards model which has an immediate effect (Approach A). The Royston-Parmar model was developed during this time and allows for a delay.

**Pre-specification rule:** Pre-specification of the statistical analysis is a rule that is routinely required in randomized clinical trials (RCTs) to prevent "the appearance that methods have been altered on the basis of the observed results to date to provide the most significant result". Therefore, the bar needs to be high before the pre-specification rule is not applied. Beyond evidence internal to the study,[18] evidence from multiple external RCTs within the field should show the original model is not correct before considering replacing the pre-specified analysis in RCTs. Furthermore, if a secondary pre-specified model is accurate, then it should be preferred over a completely new analysis as this choice lessens the appearance of altering methods to provide the most significant result. The Royston-Parmar model was pre-specified in UKCTOCS for a secondary analysis to allow for edge effects of screening. This override of the pre-specification rule applies to the proportional hazards model for the complete data from 2001 to 2020 (Approach A), because over this time the hazards are not proportional. However, for the analysis of the partial data from 2015 to 2020 (Approach B), there is much less evidence of non-proportionality, and a different contrast is needed.

**Delayed Effect of Screening:** Five RCTs for screening for cancer reported between 2009 and 2014 and all exhibited a delayed effect: (i) ERSPC for prostate cancer in 2009[14], 2012[15], 2014[16], (ii) single flexible sigmoidoscopy for colon cancer in 2010[19], 2017[19], (iii) National Lung Screening Trial (NLST) in 2011[20], (iv) C component of PLCO: two flexible sigmoidoscopies for colon cancer in 2012[21], (v) single flexible sigmoidoscopy for colon cancer in 2009[22], 2014.[23] While the power of a proportional hazards model is not significantly reduced if the delay is small compared to the trial duration, it may be significantly reduced if the delay is large, with the asymptotic relative efficiencies reduced to 30% or less for an effect delayed until half the trial time has elapsed (2$^{nd}$ of 7 non-proportionality patterns[24]) which is similar to the delay in UKCTOCS. In addition to this consistent observation, a delayed mortality effect is conceptually reasonable since early detection trials measure survival time from study entry to avoid lead-time bias. Study entry can be years prior to cancer inception, and therefore years prior to detection and subsequent treatment, leading to a delayed mortality effect. Approach C proposes a Royston-Parmar (RP) model since it allows for a delayed effect and was pre-specified in a secondary analysis. The external evidence of a consistent delay across trials of effective screening tests and the conceptual support for a delayed effect in screening, makes a case for a statistical model which allows for a delayed effect in the UKCTOCS follow-up analysis, thereby seamlessly integrating the science of early detection with the statistical thinking.[18] Changing the statistical model based on external evidence is similar to changing the endpoint of ovarian cancer, which UKCTOCS investigators judged was appropriate, from the 2003 pathology definition to the WHO 2014 pathology definition. Statistics should be allowed to employ the same scientific reasoning as pathology in allowing for updates or modifications based on new external knowledge gained during the study.

**Full vs. Partial Data Analysis:** While the delayed effect is important when analyzing the full data (2001-2020) in choosing between a proportional hazards model (Approach A) and a non-proportional hazards model (Approach C), there is no delayed effect with follow-up data where the mortality curves are already separate. Thus the choice between full data analysis (with a delayed effect) (Approach C) and a partial data analysis (Approach B) relies on different considerations, namely power and scientific consistency. Figure 1 may help in conveying the contrasts between the three approaches. The mortality curves rely on a simulation of the follow-up data based on extrapolating smooth hazard curves to 2015-2020. Since extrapolation is always uncertain the details of the model underlying the extrapolation are not important. The mortality curves continue to diverge but as noted in Approach A, the screening effect could become diluted due to ovarian cancers arising after screening ended (end of 2011) and these cases dying during the follow-up period for which there could be no effect of screening. In this situation, the mortality curves could begin to converge - thus the balance between a sustained effect of screening and dilution is impossible to predict. Because women enrolled up to 4.5 years after the study began, there is partial new data between 9.5-14 years.

The power of the approaches is mostly dependent on the difference in ovarian cancer deaths between the two arms. Approach C has greater power due to the greater area between the two curves. Approach B has almost the same area but somewhat reduced due to the progressive diminishing of weight starting at 14 years and increasing as time (censor or death) approaches 9.5 years. This progression reflects the enrollment of over 200,000 women over 4.5 years. For Approach C, but not for Approach B, the scientific description of the screening effect and the corresponding significance test are based on the same model, a scientific consistency - or a seamless integration of the science and the statistical thinking.[18]

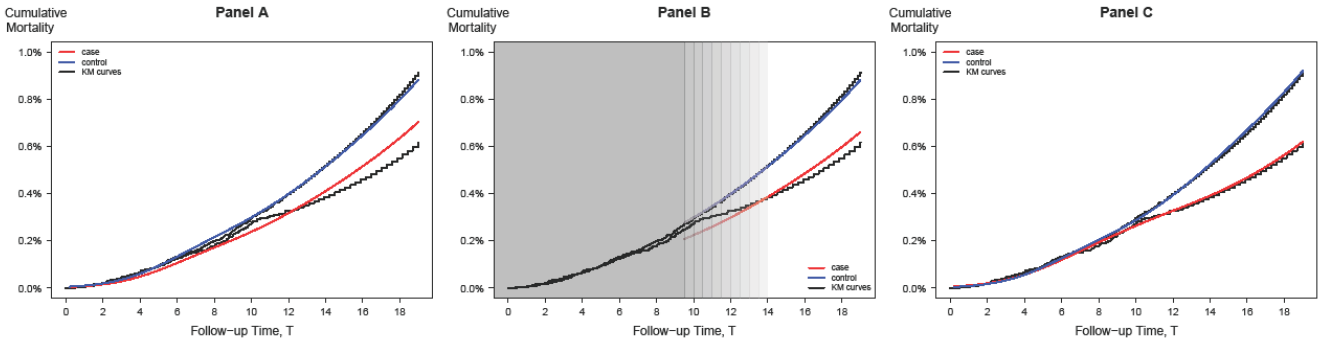

***Figure 1:*** Extrapolated hazard curves beyond 12 years yields **simulated** data beyond 12 years with *observed UKCTOCS ovarian cancer mortality curves through 12 years. Blue curves: model fit for control arm, red curves for MMS arm. Panel A displays a proportional hazards model for the full data 0-19 years* ***(Approach A)****, Panel B displays a proportional hazards model restricted to the 5 years of additional follow-up data 14-19 years with the time period (0-9.5 years) not analyzed shaded in grey, and partially analyzed data with progressive shading (9.5-14)* ***(Approach B)****, and Panel C displays a Royston-Parmar*

*model allowing for a delayed effect over the full data 0-19 years* ***(Approach C)****. Ideally the mortality curves (black) and the model (red, blue) should be close to provide an accurate scientific description, as is the case in Approach C and to some extent B. The (unshaded) area between the two fitted curves (red, blue) represents the difference in the number of OC deaths and to a large extent determines the power of the model. The greatest power (area) and the best scientific description (best fit) are provided by approach C (Panel C).*

##### 3. What are some of the potential criticisms/downside of this method?

The potential criticism of Approach C is that the standard statistical rule of pre-specification of analysis is not followed because the analysis changed from a proportional hazards model to a model that allows for a delayed effect. The argument against this is that having an analysis where the model allows for a delayed effect seamlessly integrates the science of early detection with the statistics and this approach is more important than following rules such as pre-specification as argued by David Cox.[18]

A related criticism is that we cannot 'prove' that we have not changed the form of the analysis after seeing the first (and primary) results from the trial.

#### Comment Form

| United Kingdom Collaborative Trial of Ovarian Cancer Screening (UKCTOCS) Long term follow up | | |
|---|---|---|
| **Comments from the Independent International Group on the Form of the Updated Analysis** | | |
| Question | Answer ***(please tick as appropriate)*** | **Please provide** further comments ***(as necessary)*** |
| **Which method should we use (either suggest one of the 3 proposed methods or an alternative)** | | |
| **Approach A** | ☐ | |
| **Approach B** | ☐ | |
| **Approach C** | ☐ | |
| **Alternative** (please suggest below) | ☐ | |
| **What do you see as the strengths of this method?** | | |
| **What do you see as the weaknesses of this method?** | | |

#### Supplementary Table 1 – Summary of Responses from Independent International Group

| Name | Choice of Approach | Descriptive summary of position |
|---|---|---|
| EX1 | A | “The analysis most likely to be accepted as valid by the cancer research and policy community” – uses all the data and consistent with protocol. Also suggests only include cancers diagnosed from period of intervention. Is (probably slightly) conservative. Can use C for secondary analysis. |
| EX2 | A | Keeping Cox model "avoids the appearance of trying to get a significant result by changing the test” post-hoc, having seen data. Cox/LR test requires one group to have generally larger survival times, rather than just a different distribution. Concern RP model might detect crossing/oscillating curves. Plus is RP necessarily more powerful if more df used? KM curves represent data faithfully too. Also suggest difference in crude risks (will reflect competing mortality) as clinically relevant. Acknowledges spending of α and dependent nature of LTFU. Experiment-wise p-value in 2020>0.05 |
| EX3 | A (alternative) | Suggests an alternative/hybrid of A for primary analysis and C to estimate screening effect – no post-hoc change in test (“maintain credibility in the scientific community”) but without a misleading effect size estimate. [*Note Approach A allowed other methods for complementary analyses*] OK if p-value doesn’t tally with effect size CI. Success of UKCTOCS should not be dependent on simply if p<0.05 |
| EX4 | Alternative (C?) | Recommends an alternative measure of NNS (number needed to screen) as the most suitable measure for a screening study. Says C will not be “intuitive” and RP not generally used. However, other comments suggest leaning to C in spirit: not concerned over pre-hoc versus post-hoc; concerns over multiplicity unavoidable but ‘reason’ rather than just ‘empiricism’ should have role in science. In addition, has concern about the dilution aspect – should be modelled. |
| EX5 | | *No choice indicated, but recommended a brief paper that considers long-term survival by looking at RMST from a given timepoint onwards. From email*: “This simple method does not need any modelling and the results can be interpreted easily clinically” |
| EX6 | Alternative (C?) | Has replied with a very lengthy and interesting document, discussing guiding principles first. Screening very different to therapeutic trials, patience is required – cannot expect early reductions. PHs only suitable for cancer screening when there is no benefit. Notes length-biased sampling can be factor for RCTs too – benefit more likely for slower progressing ones i.e. laters years of trial. Also questions convention of testing null, when early detection will provide at least some MR – rather, assess by how much. Generally, the method recommended is to split into yearly bins and assess HR in each, maybe with some smoothing. Avoid modelling baseline hazard – nuisance parameters and waste of df. Also consider how the results will project in practice. Single HR will give “a very blurred, incomplete and misleading picture of how much/little good screening did for the 100,000 participants, or of how much future women might expect from a screening regimen based on these screening tools.” The first HR estimate of ERSPC (=0.8) was remembered, but a larger effect in later years should have been the news. Benefits of A (continuity, appearances, simplicity) more philosophical, hard to quantify. B would give ‘disconnected’ HR, arbitrary in analysis-time. For C, there is biological (has delayed element) and statistical (has flexibility) sense but model parameters should have direct interpretation. Instead of single p-value, suggests multivariate confidence region. |
| EX7 | C | *No form filled in, but replied in email*. Generally favours C but recommends trying to find an “explicit parameterization that captures the key issues in a limited number of interpretable effects”. RP model a good empirical summary but leaves “interpretation oblique”. Ultimately though, he suggests we “do |

| | | |
|---|---|---|
| | | what you yourselves think is the most effective and secure analysis of all your data, bearing in mind the current state of information about the field, even if that is not the majority view.” |
| EX8 | C | All 3 can be tried, “but a conclusion should be reached based on a proper consideration of the full evidence” and use “scientific principles”. One of many against obsession with statistical significance. “Full information from data should be extracted”. A and B seem ‘absurd’: A appropriate under incorrect assumptions and B wastes a lot of data, plus statistically absurd *[no elaboration*]. OK to use C – have not been data-dredging or changing end-point – just “using common sense”. |
| EX9 | C | Obvious that PHs assumption does not hold in UKCTOCS (or other trials). Hence align model to reality. Main benefit of RP model is that it is informative of screening effect over time, whereas single HR applies to no time point and is hard to extract any meaningful interpretation here (main problem, not loss of power). Do A as supportive analysis only for those keen on pre-specification. B not sensible. That C was not pre-specified (except for secondary analysis) is a weakness but not a “a violation of good scientific principles”. For “a major and definitive screening trial […], such regulatory constraints should not be the primary consideration” but instead “approximating the truth as well as possible”. However, it might be better to use a model that employs fewer and more easily interpretable parameters. |
| EX10 | C (alternative) | Reason to believe PHs do not hold, so power lost with Cox. Notes that when change in HR from 1 to <1 happens after more than half events occurred, Cox is severely compromised vs composite test. B is unsatisfactory, natural to model all data. However, not convinced RP is a good idea for treatment effect test: 1) power unknown under different ‘flavours’ of non-PHs and 2) issue of simple vs complex RP model, can also affect power. Prefers to separate test from estimate, and use economical test with good power under PHs and late effects. Suggest Karrison (2016) test –“combines a standard logrank test with two weighted logrank tests, allowing for possible (near-)PH, early or late treatment effects in a single composite test.” Single WLR test not recommended as placing “eggs in the basket on a delayed effect”. Can use RP for descriptive stuff. Need to analyse survival curves respecting science; using a headline HR makes little sense if HR varies over time. Post-hoc aspect is weakness but is justifiable. |
| EX11 | C | Is not persuaded by pre-specification argument. Keeping a plan that is less preferable “turns research rules into an irrational, mindless, and restricting obsession with methodological procedure”. Experience has shown us we don’t understand the disease we are studying (and gives examples from breast cancer). “We can discern the difference in attempts by a study team to game the analysis to gain statistical significance, from a good faith effort to apply a statistical technique that is more appropriate for the data”. B opens doors to criticisms: unfavourable early results censored; early data which is important. It is necessary to understand the delayed effect. Chooses C because “rules have a purpose, but when the higher priority is understanding phenomena in a reasoned disciplined way… then a compelling argument can be made to deviate from them”. No screening trial has shown an immediate effect. RP was also pre-specified, which adds credibility. Beware ‘washout’ period.<br>Talks about how different screening trials have different results/delays, all dependent on differing facets of trial design and the cancer itself, the effects of which are largely unknown until we do the study. “Point is, we are still learning how to design and analyse RCT screening trial data.” |
